# COVID-19 endgame: from pandemic to endemic? Vaccination, reopening and evolution in a well-vaccinated population

**DOI:** 10.1101/2021.12.18.21268002

**Authors:** Elisha B. Are, Yexuan Song, Jessica E. Stockdale, Paul Tupper, Caroline Colijn

## Abstract

COVID-19 remains a major public health concern, with large resurgences even where there has been widespread uptake of vaccines. Waning immunity and the emergence of new variants will shape the long-term burden and dynamics of COVID-19. We explore the transition to the endemic state, and the endemic incidence, using a combination of modelling approaches. We compare gradual and rapid reopening and reopening at different vaccination levels. We examine how the eventual endemic state depends on the duration of immunity, the rate of importations, the efficacy of vaccines and the transmissibility. These depend on the evolution of the virus, which continues to undergo selection. Slower reopening leads to a lower peak level of incidence and fewer overall infections: as much as a 60% lower peak and a 10% lower total in some illustrative simulations; under realistic parameters, reopening when 70% of the population is vaccinated leads to a large resurgence in cases. The long-term endemic behaviour may stabilize as late as January 2023, with further waves of high incidence occurring depending on the transmissibility of the prevalent variant, duration of immunity, and antigenic drift. We find that long term endemic levels are not necessarily lower than current pandemic levels: in a population of 100,000 with representative parameter settings (Reproduction number 5, 1-year duration of immunity, vaccine efficacy at 80% and importations at 3 cases per 100K per day) there are over 100 daily incident cases in the model. The consequent burden on health care systems depends on the severity of infection in immunized or previously infected individuals.

## Introduction

COVID-19 is still spreading rapidly in many countries across the globe. There are indications that the disease will eventually become endemic rather than be eliminated. Natural questions to ask are: how will factors such as vaccination coverage, vaccine efficacy, duration of immunity and disease importation interplay to determine how and when endemic mode will be reached, and how can the transition happen without major resurgence of cases?

Despite the widespread use of highly efficacious vaccines globally, vaccines alone have failed to control transmission in many countries. Therefore physical distancing and other non-pharmaceutical interventions (NPIs) are still widely used to control the spread of COVID-19. These restrictions often come at a cost to the economy [1], and individuals’ physical and mental well-being [2, 3]. Previous studies that have investigated the impact, on COVID-19 cases, of public health measure relaxation, all agree that some level of restrictions will still be required to keep cases under control [4, 5, 6]. Since then many jurisdictions have lifted NPIs and later re-introduced them when cases surged. But at some point in the near future, it is likely they will wish to implement some level of further reopening once again. Jurisdictions will need to determine the correct level and appropriate speed of reopening to sufficiently prevent negative outcomes such as cases, hospitalizations, or deaths, in light of their vaccine uptake.

The emergence of new variants of SARS-CoV-2 virus, often called variants of concern (VOCs), is another immediate challenge for COVID-19 pandemic response. Currently identified VOCs are more transmissible than the wild-type SARS-CoV-2, and possibly have some ability to evade host immunity to COVID acquired from vaccines or infection [7, 8, 9, 10, 11, 12, 13]. SARS-CoV-2 is expected to evolve further, and new variants may continue to emerge as long as transmission remains high. Jurisdictions, therefore, need to factor in SARS-CoV-2 evolution to their COVID-19 response plan, because it has serious implications for disease control and endemicity.

It is important to study, and put in the context of a real population, how various factors such as choices related to reopening (relaxation of public health restrictions), vaccination coverage, viral evolution, waning immunity, and vaccine efficacy will shape short term case trajectories, and also determine the path from COVID-19 pandemic to endemic mode. We use two models, that we validate with a fit to data, to address several relevant issues such as SARS-CoV-2 evolution, how fast the current restrictions can be lifted without causing resurgence of cases, and the impact of high vaccination coverage on COVID-19 resurgence. We first use an age and contact structured model to assess near-term dynamics of COVID-19 under several reopening and vaccination coverage scenarios. We calculate the herd immunity threshold from an age and contact structured model, as compared to equivalent calculations in a more simple SIR model, and find good agreement. These matching estimates motivate us to develop a simple SVEIRS model to investigate how factors including vaccination efficacy against infection, infection importation rate, waning rate of acquired immunity, and the emergence of high transmission variants will impact the endemic state of COVID-19. The simpler model allows us to obtain a closed-form solution at the endemic steady state, and predict as well as analyze case incidence at endemicity. We also explore how antigenic drift and shift compare in this model, in terms of reduction in vaccine efficacy and the resulting impact on COVID-19 case numbers.

## Methods

In this study we use two Susceptible-Exposed-Infectious-Recovered models to answer important public health questions about the impact on the path to COVID-19 endemicity of vaccination coverage, public health measure relaxation plans, viral evolution, and immunity waning rates. The first model is an age and contact structured deterministic model. The second, simpler, model is a classic SEIRS model with vaccination. Both models are set up to reflect the pandemic trajectory in the British Columbia, Canada (BC) population of just over 5 million people. We present results as a rate per 100K population for infections and hospitalizations. Model codes and data for both models are openly available in: https://github.com/Yexuan-Song/End-Game.git

### Model 1: Age and contact structured model

This is a Susceptible-Exposed-Infectious-Recovered model in which the population is stratified into 15 sub-populations: by age {0–9, 10–19, 20–29, …, 70–79, 80+} and work status {20 *−* 29^*e*^, 30 *−* 39^*e*^, …, 70 *−* 79^*e*^}. Groups with superscript *e* denote an “essential worker” group. The model has a contact matrix that represents the contact probability between each age and “essential worker” status group. The reproduction number *R*_NPI_ reflects the effective reproductive number in the absence of vaccination but in the presence of NPIs (social distancing, quarantine, school opening etc.). The model tracks vaccination status, but only whether an individual takes a vaccine or not, neglecting the details of number of doses and time until the vaccine is effective. There is no waning of acquired immunity in the model, and we assume that vaccines can still prevent hospitalization and death even when they fail to prevent infection. We therefore use this model to explore short-to-medium term dynamics of COVID-19, primarily the impact of relaxation of NPIs and of vaccine coverage. We model gradual reopening (relaxation of NPIs) by increasing *R*_NPI_ in a linear fashion from *R*_NPI_ = 2.2 over a 300 day window, while rapid reopening is modelled by an all-at-one increase of *R*_NPI_. This age and contact structured model was presented in [5], and is based on an earlier model of [14].

#### Rationale for *R*_NPI_

Estimates of the reproductive number for COVID-19 in the absence of social distancing restrictions range from 2 to 4 for non-VOC SARS-CoV-2 [15]. The delta variant has a higher transmission rate than previously-predominant strains, which acts to increase *R*_NPI_. It has also been reported to show some potential for immune escape [13]. However, we model the maintenance of symptomatic testing and contact tracing, which can reduce *R*_NPI_ by 1/3 if done rapidly, optimally, and with the capacity to scale up as cases rise [16]. We therefore explore relaxation from *R*_NPI_ = 2.2 to *R*_NPI_ = 2.5, both in the rapid and gradual reopening scenarios. In supplementary analyses, we explore relaxation to a wider range of *R*_NPI_ values, ranging from 2.4 to 5. Note that in British Columbia, early estimates of *R*_0_ for the original COVID-19 virus were approximately 3 [17]; with the Alpha and Delta variants both estimated to increase the transmissibility by approximately 100%, this would place Delta (currently predominant) at an *R*_0_ of over 6 [18, 19]. However, a range of measures are in place at the time of writing, including testing, indoor mask use and vaccine mandates, workplace screening and other measures, motivating a lower *R*_NPI_.

#### Vaccine efficacy

In our model vaccine efficacy consists of two components: *v*_*e*_, efficacy against infection (what fraction of infections are prevented) and *v*_*p*_, efficacy against symptoms when infection does occur (what fraction of cases infected after vaccination do not have symptoms, including severe outcomes). To have a symptomatic case after vaccination, the vaccine has to both fail to prevent disease and symptoms, so efficacy against symptomatic infection is *v*_*d*_ = 1 − (1 − *v*_*e*_)(1 − *v*_*p*_). We take our baseline values from studies on the Pfizer vaccine, giving *v*_*d*_ = 95% protective against symptomatic infection [20] and *v*_*e*_ = 80% protective against infection [21], implying a value of *v*_*p*_ = 75%.

If the vaccine fails to protect against infection in an individual, but does prevent symptoms, in our model the individual is assumed to contract the virus and transmit to others at the same rate as an unvaccinated individual. It is likely that those who are vaccinated but infected anyway are less infectious due to lower viral loads than those who are infected without vaccination, and it is likely that they would not have symptoms. In a framework where testing is driven by symptoms and those with symptoms are encouraged to isolate, asymptomatic individuals will not know they are ill and will likely remain circulating and infectious for longer than those who develop symptoms. Thus, longer duration of infectiousness (vaccinated-but-infected individuals may transmit more because they do not have symptoms and therefore do not isolate), and lower per-unit-time transmission (due to a reduced viral load) act in opposing directions. In our model we assume that these effects balance out.

#### Vaccine acceptance

We model age-based vaccine uptake according to data from the vaccine rollout by age in BC [22]. This includes highest uptake in older age categories (98% in those 80+), and lowest uptake in younger age categories (30% in those 12-19). Vaccine hesitancy is modelled as around 10% lower in essential worker categories. A full description of the model age-based vaccine uptake is included in Supplementary Table S1.

#### Vaccine rollout

Following BC vaccine rollout strategy [22], we vaccinate those 80+ and in long term care (LTC) settings (not modelled explicitly, but hospitalization rates are adjusted to reflect protection in LTC [5]) first, followed by younger age groups 12+. To match the observed age-based trajectory [22] (whilst taking into account that we do not model the two-dose vaccine regimen), we model a rapid uptake in vaccines during May-June 2021 in which 2% of each age group are vaccinated per day up to uptake levels observed during that time (see Supplementary Table S1). This is followed by a slower period of 0.2% per day in July-Aug and then 0.22% per day from September onwards, until the overall uptake levels described in in Supplementary Table S1 are reached. Vaccination of those aged 12-19 does not begin until July 3rd 2021. The full age-based rollout is shown in Supplementary Figure S10.

#### Model Validation

We validate the age and contact structured model by matching the model predicted case counts (including vaccine uptake and rollout as detailed above) to reported cases by age in BC from January to November 2021 (Figure 1).

**Figure 1:**
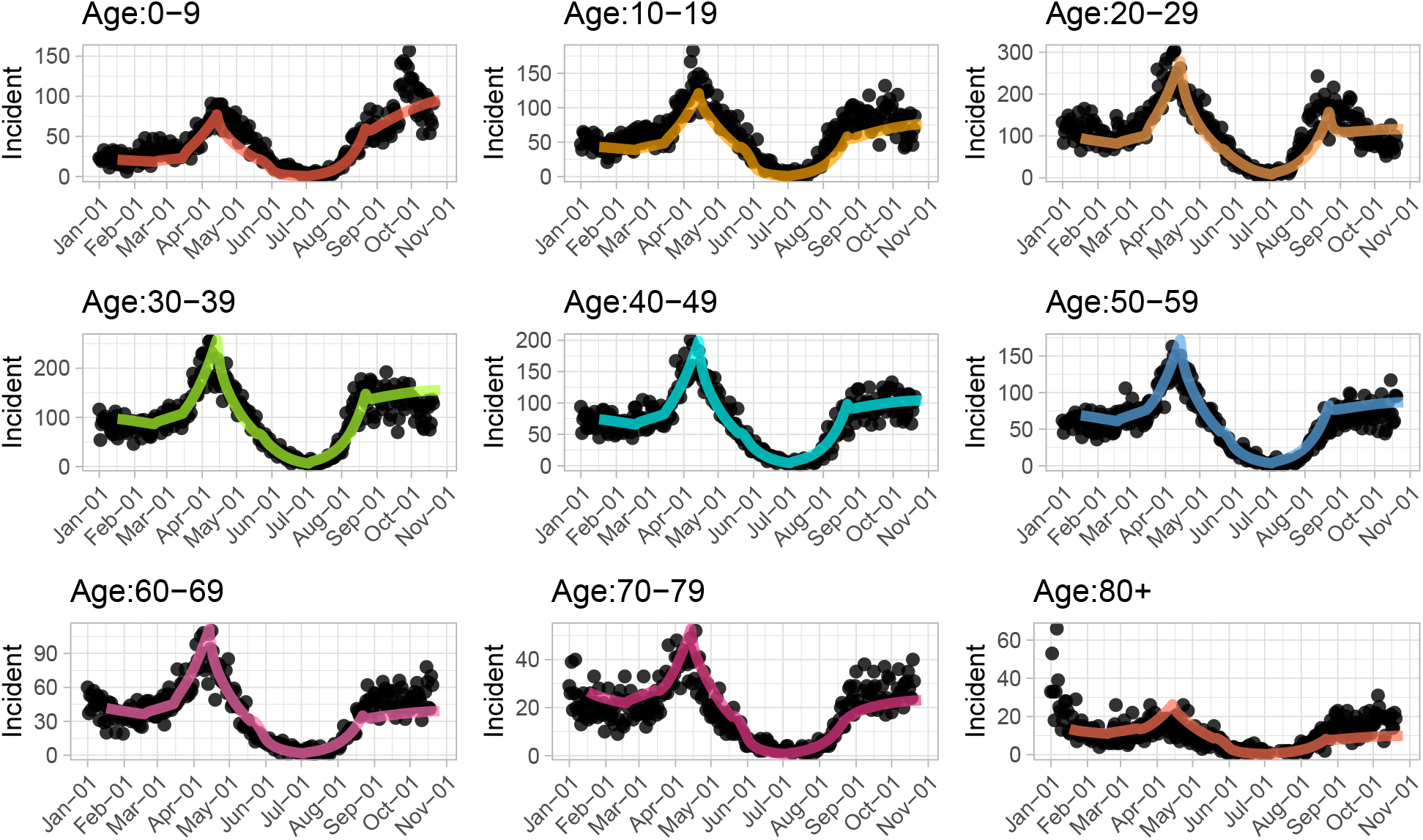
Model validation with age distribution of cases. Black dots represent actual reported cases by age from January 2021 to November 2021 in British Columbia, Canada; colored lines are the model predicted case counts by age.

On these dates, the model contact matrix and reproduction number are modified to fit the reported case counts by age. A list of the identified important dates is included in Supplementary Table S2. As shown in Figure 1, the model output matches reasonably well the reported case numbers.

### Model 2: Exploring endemic state with a simple SVEIRS model

Key determinants of the endemic state of COVID-19 pandemic are: viral evolution — which will determine the overall transmissibility of infection, and also dictate the antigenic drift and/or shift of the virus over time, infection importation rates, vaccine uptake and vaccine’s efficacy at preventing infection, as well as the duration of acquired immunity. To investigate how these factors interplay in determining the path to COVID-19 endemicity, we develop a simple Susceptible-Vaccinated-Exposed-Infectious-Recovered-Susceptible compartmental model to analyse and predict the endemic state for the COVID-19 pandemic in BC. The model is described with a system of first order ordinary differential equations (see below). In the model, susceptible individuals are vaccinated at a rate *ν* per day, and we assume *v*_*e*_ vaccine efficacy against infection. Both vaccine induced and infection induced immunity wane at a rate *w* = 1/*D*, where *D* is the duration of acquired immunity. A quantity *c*(*t*) models control interventions at time *t* by regulating the transmission rate *β* to reflect changes in disease transmission when measures are implemented or relaxed in response to either resurgence of cases or to ease the negative impact of lockdown restrictions when cases are deemed to be under control. Exposed individuals become infectious after an average 1/*σ* days, and they eventually recover, or are removed, after an average 1/*γ* days. To model importations, the model allows *f* susceptibles per unit time to be replaced by *f* individuals who have already been infected but are not yet symptomatic or infectious, modelling the impact of travel-associated introductions without net changes in the population size. We use the following baseline parameter values: *ν* = 0.7% per day, *γ* = 1/6, and *σ* = 1/3[23]. Note that *ν* is the per day rate for renewing vaccination (through boosters, for example) *after* individuals’ immunity has waned. We explore several scenarios that could determine the path of transition of COVID-19 pandemic to its endemicity in BC.

The model equations are as follows:

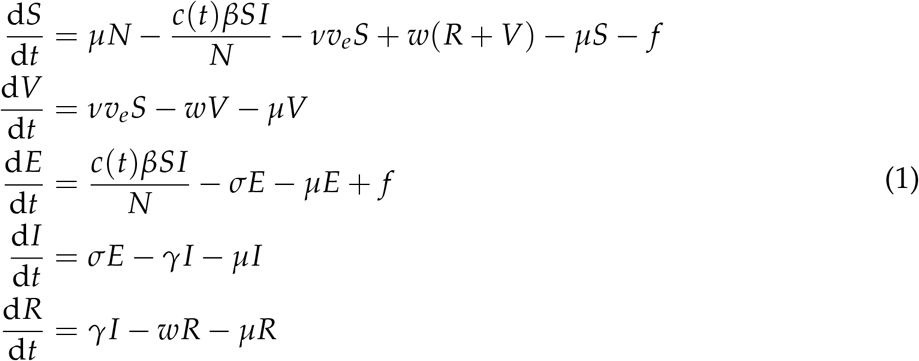

where *N* = *S* + *V* + *E* + *I* + *R*.

The prevalence (*I*\*) at endemic steady state is obtained analytically and analysed as a function of various parameters representing the aforementioned factors that will determine the number of active cases when the disease becomes endemic. For simplicity, we assume that *c*(*t*) = 1 at endemic state (no physical distancing measures, but we explore a range of transmission parameters) and that the population is constant over time. The full analytic solution is provided in the Supporting Information.

#### Model Validation

Model validation involves matching COVID-19 reported cases, from February to November 23, 2021, to model outputs. The vaccination rate was set such that the vaccination coverage in the model largely resembles vaccination uptake in BC during that period. The model fit to data is shown in Figure 4.

## Results

We explore the severity and risk breakdown of gradually reopening over a 300 day period compared with a near-instantaneous reopening, using the age and contact structured model (Figure 2). We find that under our baseline vaccine efficacy assumptions, even after most of the rollout is complete, we will not be in a position to reopen without seeing a rise in cases. This is consistent with the fact that the current decline in cases (approximately 2% per day) is slow, leaving little leeway for increasing transmission without moving from a decline to a rise. From the date of reopening (Dec 2021) onwards, near-instantaneous reopening leads to 500K cumulative reported cases and 19K hospitalizations over 900 days while gradual reopening leads to 450K cumulative cases and 17K hospitalizations, in the BC population of 5 million people. Further scenarios are explored in Supplementary Figures S4, S5 and S6, where we consider gradual vs rapid reopening from *R*_NPI_ = 2.2 to *R*_NPI_ = 2.4, 2.6 and 3.0, respectively. In all scenarios considered, there are fewer cumulative cases and hospitalizations under gradual reopening than under rapid reopening.

**Figure 2:**
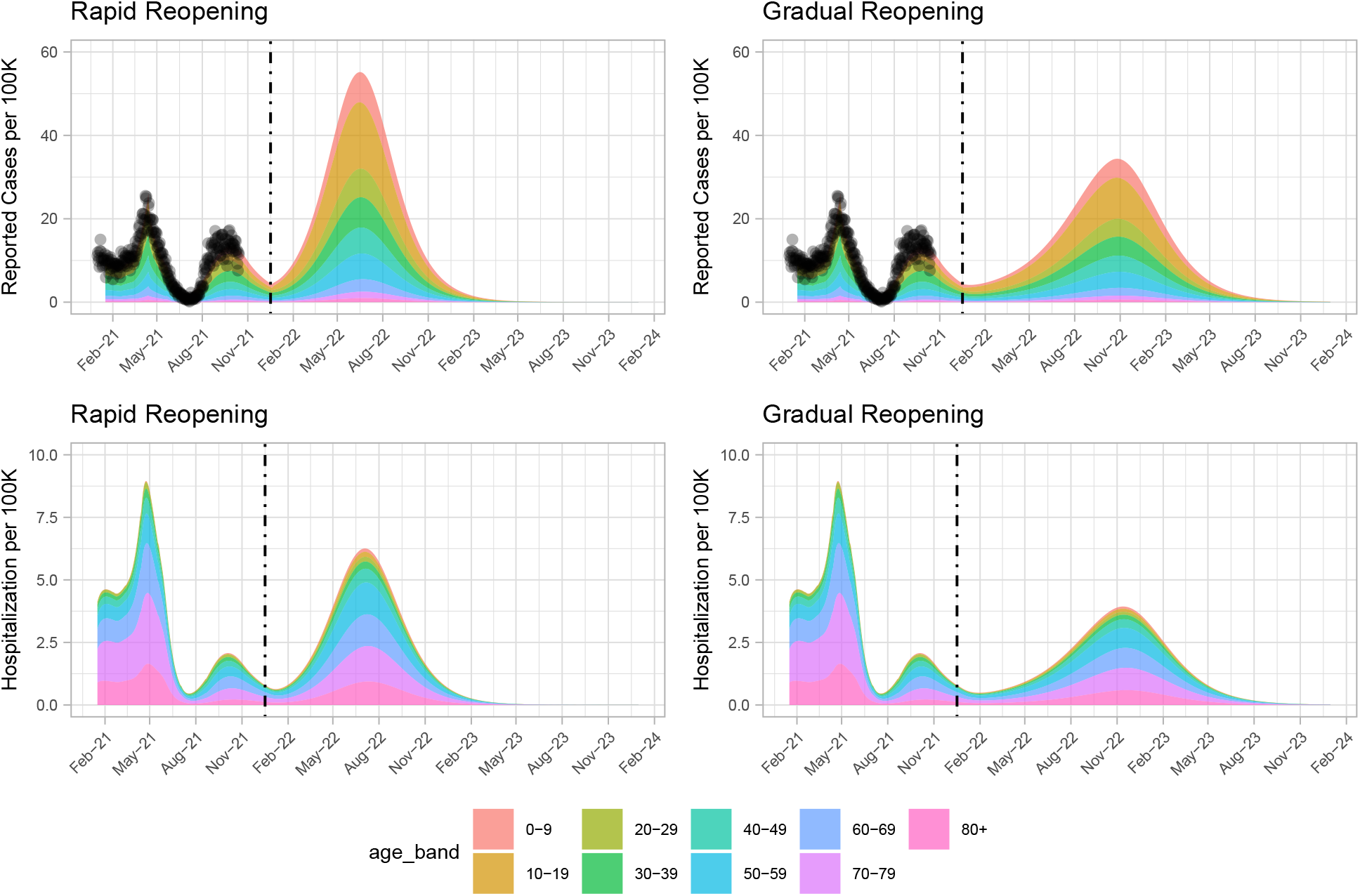
Comparison of the impact of gradual vs rapid reopening, from *R*_NPI_ = 2.2 to *R*_NPI_ = 2.5, on case and hospitalization rates. Gradual reopening is modelled as occurring linearly over a 300 day window. We assume 90% vaccination coverage of those aged 12+ before reopening, consistent with BC at the time of writing. Dashed lines represent reopening time. No further vaccines are deployed after reopening in both scenarios.

We considered reopening at two levels of vaccination coverage - BC’s 90% coverage and a counterfactual 70% scenario. Figure 3 shows the simulated outcome if we relax measures to *R*_NPI_ = 2.5 for the two scenarios, according to the age and contact based rollout. Reopening “fully” to *R*_NPI_ = 2.5 at 70% vaccination coverage leads to considerable rises in infections and hospitalizations. In both cases hospitalizations do not exceed hospital capacity (just under 40 per 100K) [24], with estimated maximum hospitalizations at 20 and 9 per 100K, for 70% vaccination and 90% vaccination, respectively. However, reopening when 70% are vaccinated leads to far more hospitalizations (600K cumulatively) than when reopening after 90% of the population are vaccinated (450K cumulatively). Further scenarios are shown in Supplementary Figures S7, S8 and S9, where we consider reopening to *R*_NPI_ = 3.0, 4.0 and 5.0, respectively, and show the cumulative cases for each scenario. 90% vaccination coverage naturally leads to substantially fewer cases than 70% coverage.

**Figure 3:**
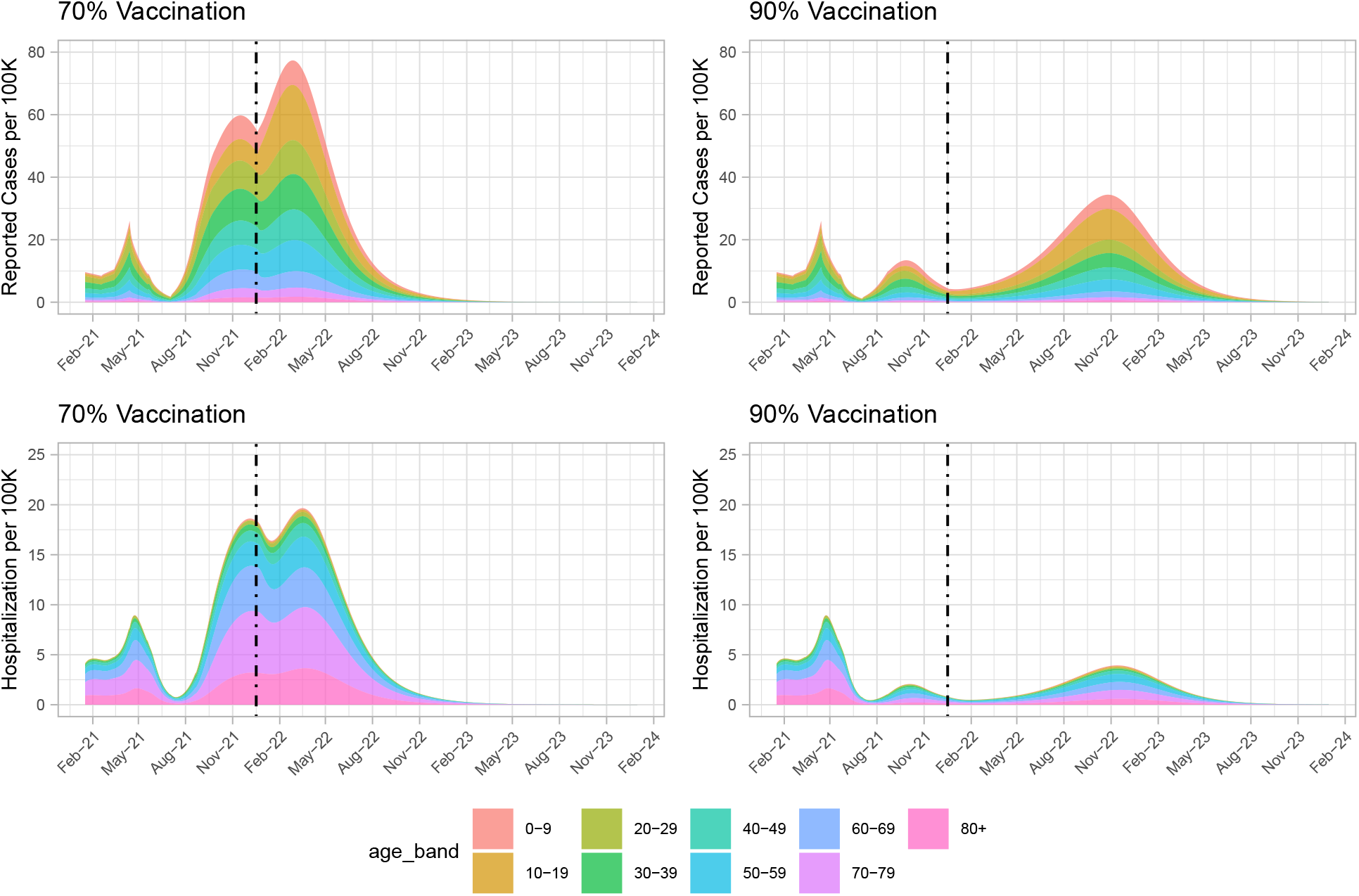
Reopen gradually at 70% or 90% vaccination coverage. Left frame showing reported cases per 100K of the population for the 70% vaccination coverage scenarios. The right panel reported cases per 100K of the population for the 90% vaccination coverage scenarios. Assuming reopening to *R*_NPI_ = 2.5.

We compare the age and contact structured model’s results to those obtained from a theoretical (SIR) model of herd immunity. Figure S1 illustrates that the age and contact structured model’s prediction for the fraction of the population protected at the herd immunity threshold is similar to the theoretical prediction from the simple model. We can therefore estimate whether a given level of immunity obtained through vaccination is sufficient to stop the spread of COVID-19 using the classic relationship in SIR models between minimum herd immunity fraction *f*_*m*_ and reproductive number *R*_0_: *f*_*m*_ = 1 − 1/*R*_0_. Consider this simple theoretical example: in a jurisdiction where 20% of the population declines the vaccine, 10% are not eligible and we have a vaccine that is 80% effective against infection, the fraction of the population that is immune from vaccination alone is *f*_*m*_ = (1 − 0.2)(1 − 0.1)0.8 = 57.6%. The 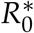 to which that fraction confers herd immunity, 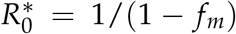, is then 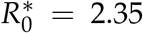. Accounting for approximately 5% of the population having had COVID-19 but some overlap between past infection and vaccination, *f*_*m*_ ≈ 60%, with corresponding 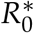 of 2.5. Higher *R*_0_ values lead to rises in cases.

Given the motivation of similar endemic behaviour in the age and contact structured model and the SIR model above, we continue to explore several long-term scenarios using the SVEIRS model, which includes waning immunity. We first consider reopening to various *R*_NPI_ values, whilst also allowing for importation of infected cases. Our simpler model results suggest that there may be multiple waves of COVID-19 cases before it eventually becomes endemic. The frequency and peaks of the waves will depend on the duration of immunity and whether or not the vaccination campaign will continue to be supplemented with booster doses. When *R*_NPI_ = 5 or greater, cases rebound very quickly to cause another major wave. In contrast, if *R*_NPI_ is below 4, reopening will not lead to a major wave before becoming endemic. This is under the assumption that booster doses will be used to maintain relatively high population immunity (Figure 4 A). Furthermore, we study several immunity waning regimes (Figure 4 B). The endemic state is sensitive to the duration of acquired immunity, even under continual boosting after immunity wanes. Reopening to *R*_NPI_ = 3.5 where immunity lasts for 1 year will lead to gradual resurgence of cases, with high endemic incidence close to 40 reported cases per 100K per day. The picture becomes more optimistic as immunity lasts longer (Figure 4 B), with endemic incidence closer to 5 reported cases per 100K per day under 2-3 year immunity. However, if booster doses are suspended and immunity wanes, the projections become very pessimistic (See Figure S2 in the Supporting Information). This will be compounded if high transmission and immune escape variants continue to emerge.

**Figure 4:**
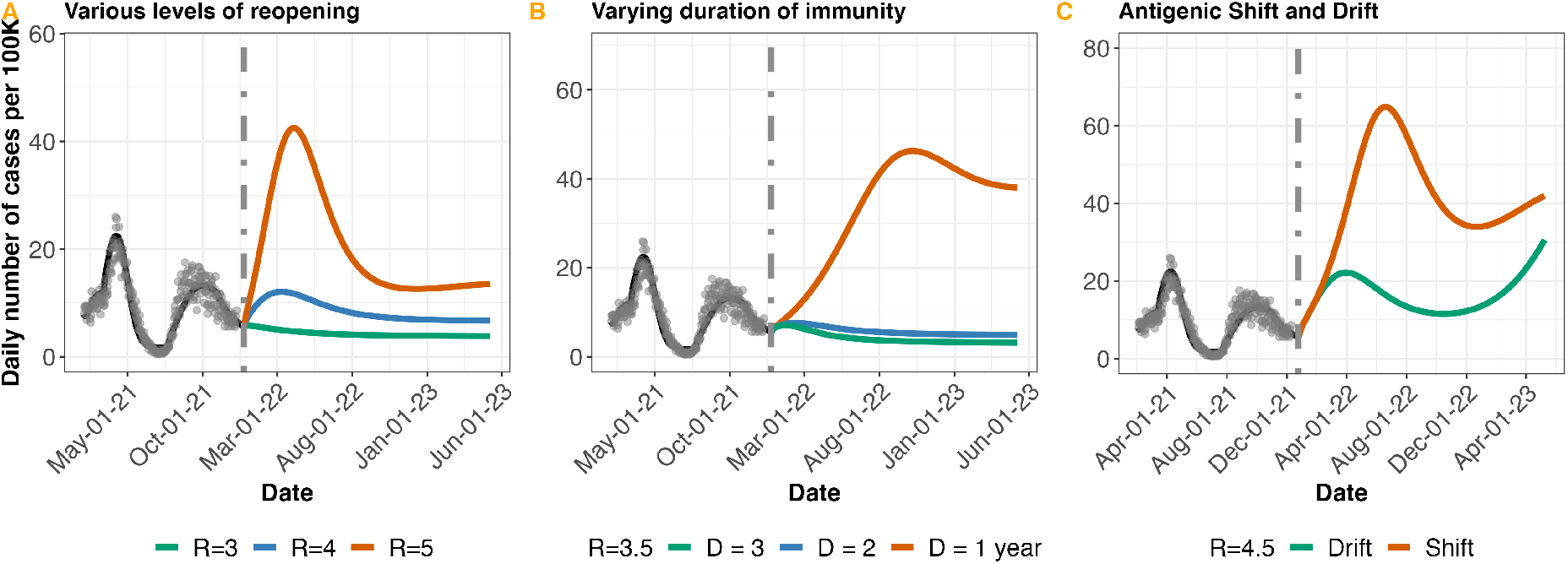
Near-future model projections and various possible paths to COVID-19 endemicity in BC. A. Projected daily cases for different levels of reopening (shown by changing R), assuming reopening occurs end of November 2021. B. Projected daily cases for different lengths of duration of immunity (1–3 years), at *R*_NPI_ = 3.5. C. Comparing antigenic drift and shift. Gradual decrease in vaccine efficacy (“drift”) over a 500-day period versus a sudden decrease (“shift”) from 80% to 40% over a few days, at *R*_NPI_ = 4.5. Model output is matched to reported cases (grey dots) in BC from February to November 23, 2021. Dotted vertical line indicates further reopening. Where not varied, importation rate *f* is fixed at 2 cases per 100K per day, vaccine efficacy *v*_*e*_ at 80%, and duration of immunity *D* at 2 years.

We compare the impact on COVID-19 dynamics of gradual changes (or small mutations) in the virus that make vaccines less effective against them over time, compared to more abrupt mutation(s) that reduce vaccine efficacy more rapidly. Borrowing terminology from influenza viruses, we term these “drift” and “shift”, though the biological mechanisms will differ. One rationale for considering lower efficacy is the continued emergence and spread of VOCs that may undermine vaccination as a COVID-19 control strategy. At the current time, evidence suggests that antibody neutralization is not as effective for VOC B.1.351 [25] and P.1 [26] as it is for the SARS-CoV-2 variants we have seen to date (including B.1.1.7) — although vaccines’ population-level effectiveness against VOCs are still relatively high [27]. At the time of writing the extent to which the rapidly-rising Omicron variant evades immunity is unclear, but it shows reduced antibody neutralization [28, 29]. We model “drift” and “shift” by reducing vaccine efficacy *ν*_*e*_ gradually from 80% to 40% over a 500-day period, and a sudden (all-at-once) change, respectively. We find that a sudden shift leads to a worse outcome in the model, with a steep rise and fall before the system settles to endemic equilibrium (Figure 4 C).

We explore the impact of four endemicity-determining factors on the endemic incidence: reproduction number, immunity duration, vaccine efficacy and importation rate (Figure (5) A to F). The endemic incidence is sensitive to all of these unknown factors, but is most sensitive to the combination of the underlying transmission *R*_NPI_, vaccine efficacy and the duration of immunity. The model’s endemic incidence is not always markedly lower than peak incidence levels in the pandemic to date (approximately 20 per 100K per day). High endemic levels occur if immunity wanes rapidly (in under 1.5 years), if *R*_NPI_ for the combination of virus and long-term measures is above 3 if there are over 6 imported infections per 100K per day, if efficacy is low and for various combinations. We model the true incidence; reported incidence would be lower, and would depend on the surveillance system that is in place and on the extent to which infection caused symptoms and severe disease.

**Figure 5:**
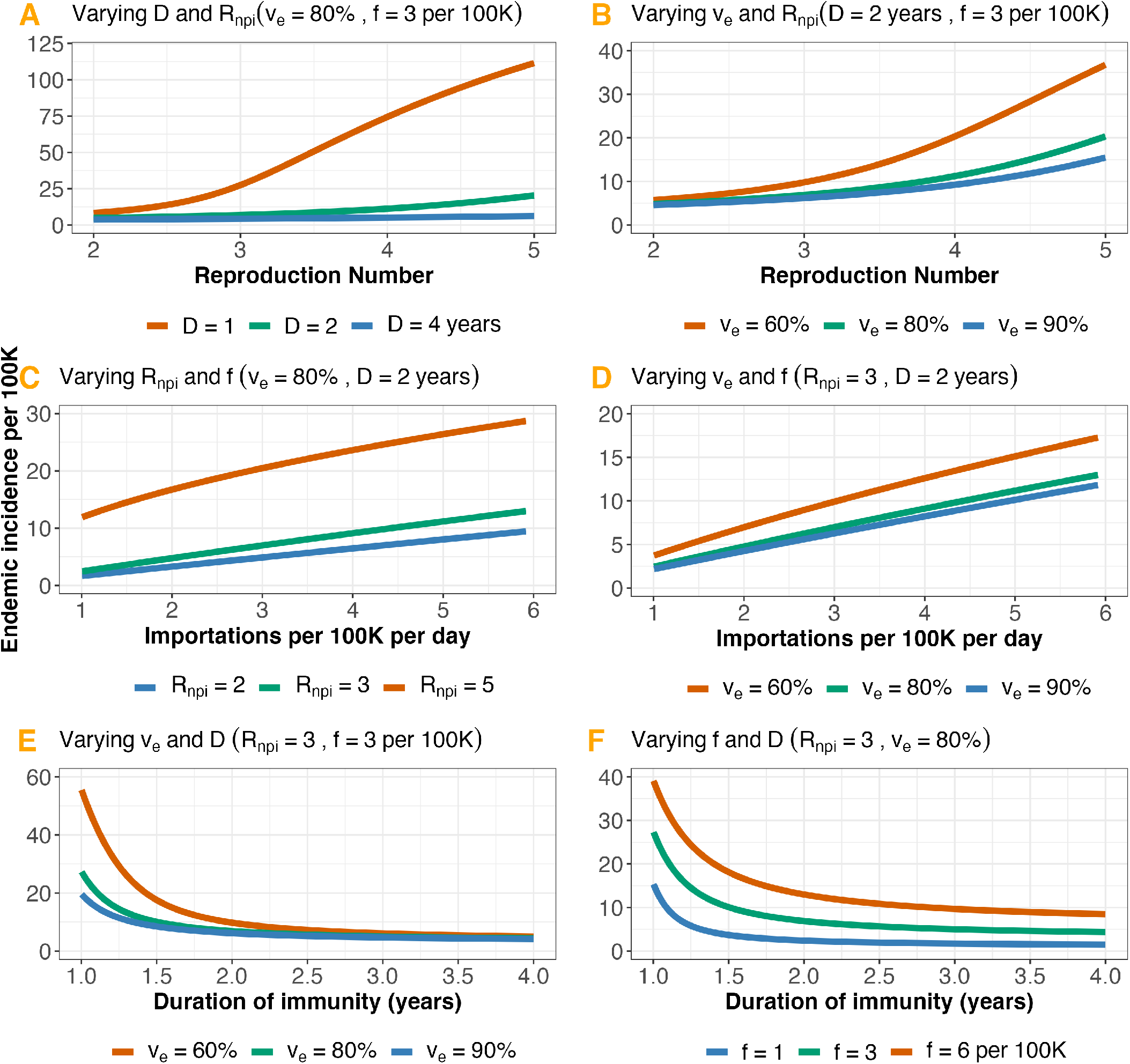
Endemic incidence as a function of: A. The reproduction number (*R*_NPI_) and duration of immunity (*D*). B. *R*_NPI_ and vaccine efficacy (*v*_*e*_). C. Importation rate (*f*) and *R*_NPI_. D. *f* and *v*_*e*_. E. *D* and *v*_*e*_. F. *D* and *f*. We set baseline parameter values such that vaccine induced immunity and immunity due to infection last for 2 years, and boosters are given 4 months after immunity wanes. Parameter values are: *f* = 3 cases per 100K per day, vaccination rate *ν* = 0.7% per day, *D* = 2 years, *v*_*e*_ = 80%, and *R*_NPI_ = 3. Numbers are not adjusted for ascertainment.

We note that when *R*_NPI_ is relatively low and vaccine coverage is substantially high, our model predicts no incident cases without importations, and in this sense it is an optimistic baseline from which to explore. In practice, heterogeneity in the population, introductions from animal reservoirs, continued viral evolution and other factors not included would likely mean that instead there would be some very low level of endemic incidence at our baseline parameters.

## Discussion

Our results suggest that COVID-19 cases will rebound after restrictions are lifted completely under current vaccination rollout plans and vaccine efficacy. In all of our models, children, adults who decline the vaccine, and adults for whom the vaccine did not prevent infection are numerous enough that the pandemic can unfold among them once restrictions are lifted. This occurs even under optimistic assumptions that immunity is continuously boosted, that those who have recovered are not at all susceptible to reinfection, and that VOCs for which vaccines are less effective than current estimates for the mRNA vaccines are not circulating, as are limitations in our age and contact structured model. Some of our model estimates of endemic incidence are similar to the peak incidence observed in BC during the pandemic so far. On the path between the current state of the pandemic and the eventual endemic state, the speed and peak of case resurgence will be modulated by how fast we reopen, vaccination coverage and vaccine efficacy, as well as the transmissibility and immune escape capacity of the dominant variant at the time of reopening.

At the time of writing, many EU countries are experiencing surges in COVID-19 cases, after many of the countries reopened at 70% vaccine coverage. Large resurgences are to be expected under those circumstances, because vaccine effectiveness is not 100% and the transmission rate of the Delta variant is very high. In Austria, for instance, where only 65% [30] of the population are fully vaccinated, daily cases are at all time high with more than 15K reported cases per day. Austria has now reintroduced lockdown restrictions for unvaccinated to curb resurgence in COVID-19 cases.

Currently, vaccine effectiveness (while not 100%) is high against infection and disease. However, the SARS-CoV-2 virus is only newly facing large vaccinated populations, and has primarily experienced selection in favour of enhanced transmission to date [31]. Indeed, this selection played a role in the rapid emergence of several VOC including both and P.1 [31]. However, as populations across the world become vaccinated, SARS-CoV-2 will face increased selection in favour of immune escape, and decreased selection for higher transmissibility. SARS-CoV-2 remains a relatively new virus, and we should anticipate that it will evolve further. Accordingly, Delta and other currently-known VOC will not likely remain the key threats to vaccination’s effectiveness in ending the pandemic. We found that sudden shifts in efficacy are more dangerous than slower drift, in causing significant setbacks in the COVID-19 response. The sudden emergence of the Omicron variant (B.1.1.529) which has 32 mutations in the spike protein along with mutations in other regions of the genome (see preliminary data [32]) may indicate that SARS-Cov-2 is capable of sudden changes in immune evasion. Similar patterns have been observed for type A influenza viruses, which experience both antigenic shift and drift and are more likely to cause major outbreaks than type B viruses that experience only antigenic drift [33].

The large peaks predicted by the simpler model may not be observed, because it is unlikely that cases will be allowed to grow excessively before some public health measures are taken. NPIs such as travel restrictions or physical distancing measures could be reimplemented to control rising cases. Booster doses could further reduce population susceptibility [34]. On the other hand, there may be limited motivation to curb transmission if SARS-CoV-2 ultimately presents as mild disease in most people, for example due to cross immunity and/or residual immunity from vaccination or previous exposure [35]. If measures are not implemented, the predicted high peaks could be observed, particularly if testing rates are high. We assumed constant ascertainment over time, but ascertainment rates can change rapidly depending on testing criteria, test-seeking behaviour, and likelihood of symptoms and severe disease.

The endemic prevalence of infection will determine the endemic demand for hospital and acute care resources, though both ascertainment and the relationship between infections and hospitalizations may change. Eventually, nearly all infections will occur in those who were either immunized or previously exposed, and with B-cell mediated immunological memory that is long-lasting, it is to be hoped that 100 per 100K incidence (Figure 5 A) will not present a burden to the health care system so strong as to require widespread NPI measures. However, throughout the pandemic in BC to date, reported COVID-19 cases have been hospitalized at a relatively constant rate around 9% [36]. This rate has been largely unaffected by vaccination. Early observations suggest that disease-blocking immunity wanes more slowly than infection-blocking immunity [35, 37]. If this is the case, we can expect the rate at which cases are hospitalized to decrease at endemic state. However, if the endemic incidence is high (over 30 incident infections per 100K per day), even a large reduction in overall severity (such as an 80% reduction) would leave on average just under 30 daily hospitalizations. Current conditions suggest that this would place a burden on the health care system, particularly if it were enhanced by seasonal variation, and if capacity were impacted by other seasonal infections such as influenza.

Overall, the virus’ evolution and the nature of waning immunity will shape the relationships between infections and reported cases, and between infections and hospitalizations/health care burden. If immunity against infection wanes quickly while immunity against disease lasts longer, and testing criteria are largely symptom-based, then reported cases might be low even where there are ample infections, presenting the opportunity for immune-evading variants to emerge. Population-level screening and genomic surveillance will aid in the rapid detection of emerging types and the assessment of their phenotypes.

This study shows that the endemic mode can be reached without risking resurgence of cases, if restrictions are lifted slowly, and measures are taken to increase vaccine uptake, while closely monitoring disease importations and viral evolution to enable quick detection/identification of VOCs. However, without carefully planned and properly executed interventions, COVID-19 may continue to cause considerable public health disruption for several years to come.

## Data Availability

All data produced are available online at https://github.com/Yexuan-Song/End-Game.git

## Funding Acknowledgement

EBA, YS, JES, and CC are funded by the Federal Government of Canada’s Canada 150 Research Chair program. PT and CC were supported by Natural Science and Engineering Research Council (Canada) Discovery Grants (RGPIN-2019-06911 and RGPIN-2019-06624).

## Supporting Information

The prevalence at endemic equilibrium is obtained analytically by solving equation (1) at equilibrium.

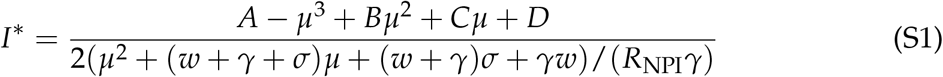

where:

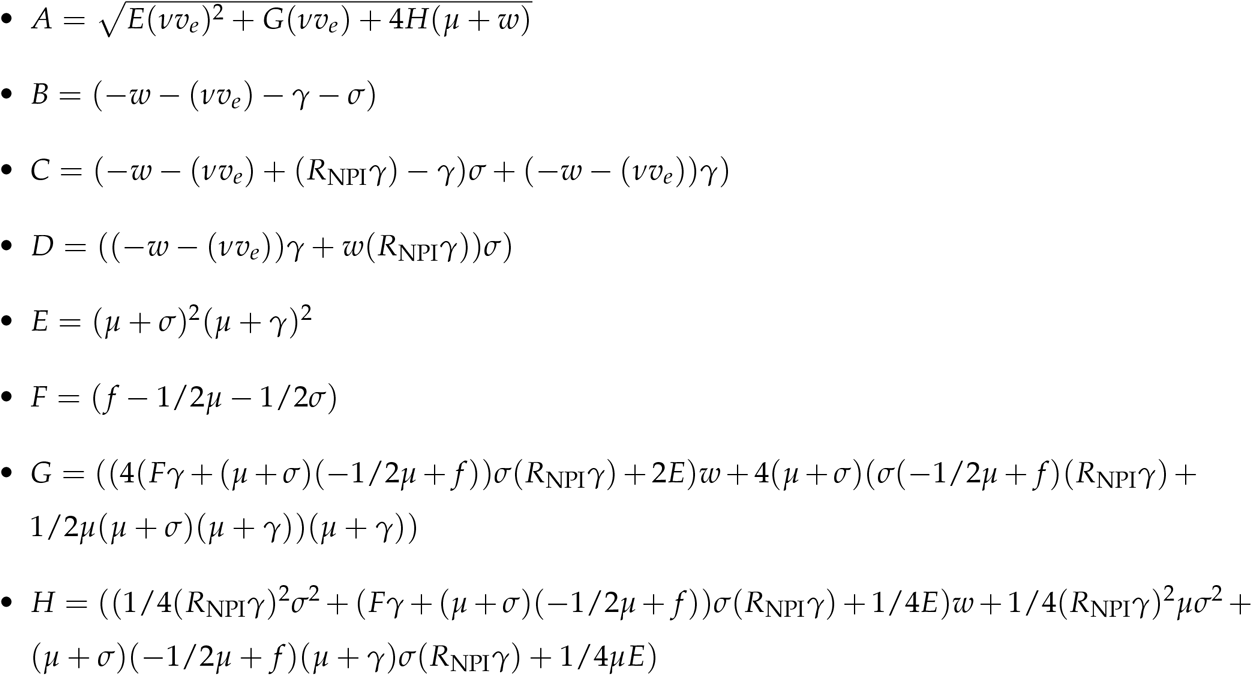

Equation (1) has no disease free equilibrium when *f >* 0, since cases are continuously introduced into the population. When *f* = 0, the disease free and endemic equilibrium points exist and their stability depends on *R*_NPI_. The reproduction number in the absence of vaccination is 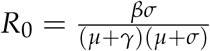, with vaccination and waning 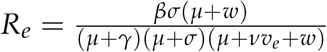 We compare herd immunity threshold calculated from the age and contact structured model and a simple SIR model, and we find good agreement (Figure S1). Also, we explore scenarios where booster doses are not given after 70% of the population are immune to infection due to vaccination. The results are shown in Figure S2.

**Figure S1:**
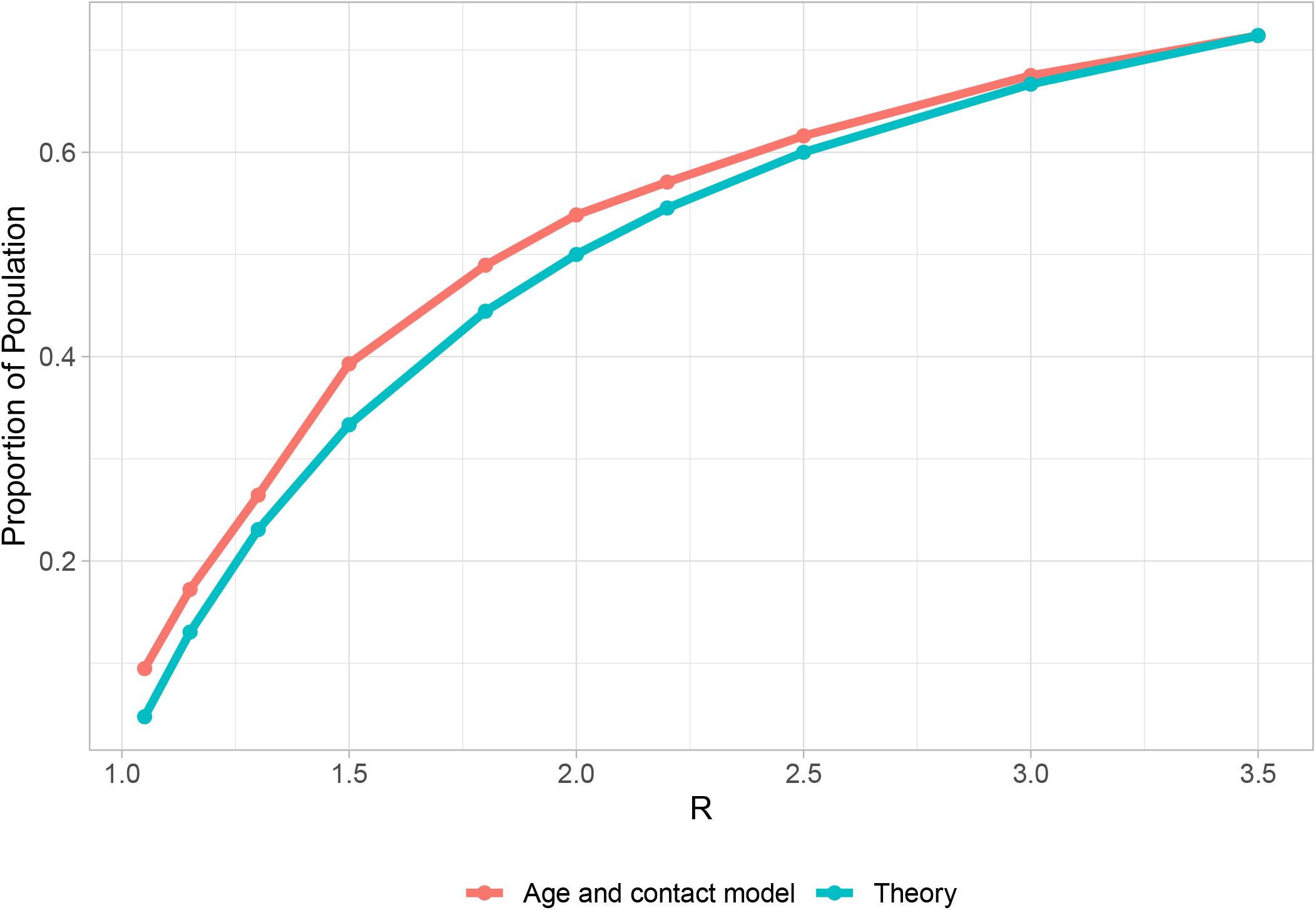
Comparison between herd immunity estimates from age and contact structured model and a simple SIR model. The age and contact structured model is very similar to a simple SIR model in terms of the fraction of the population that must either be infected naturally (in the *E, I* or *R* classes) or vaccinated successfully in order for the number of infections to begin to decline. This is the so-called “herd immunity” fraction. The theoretical result (blue) is simply 1 − 1/*R*. The model result is obtained by running a simulation at the given *R*_NPI_, as always defined in the absence of vaccination, detecting when infections begin to decline, and obtaining the portion of the population either infected or successfully vaccinated at that time.

**Figure S2:**
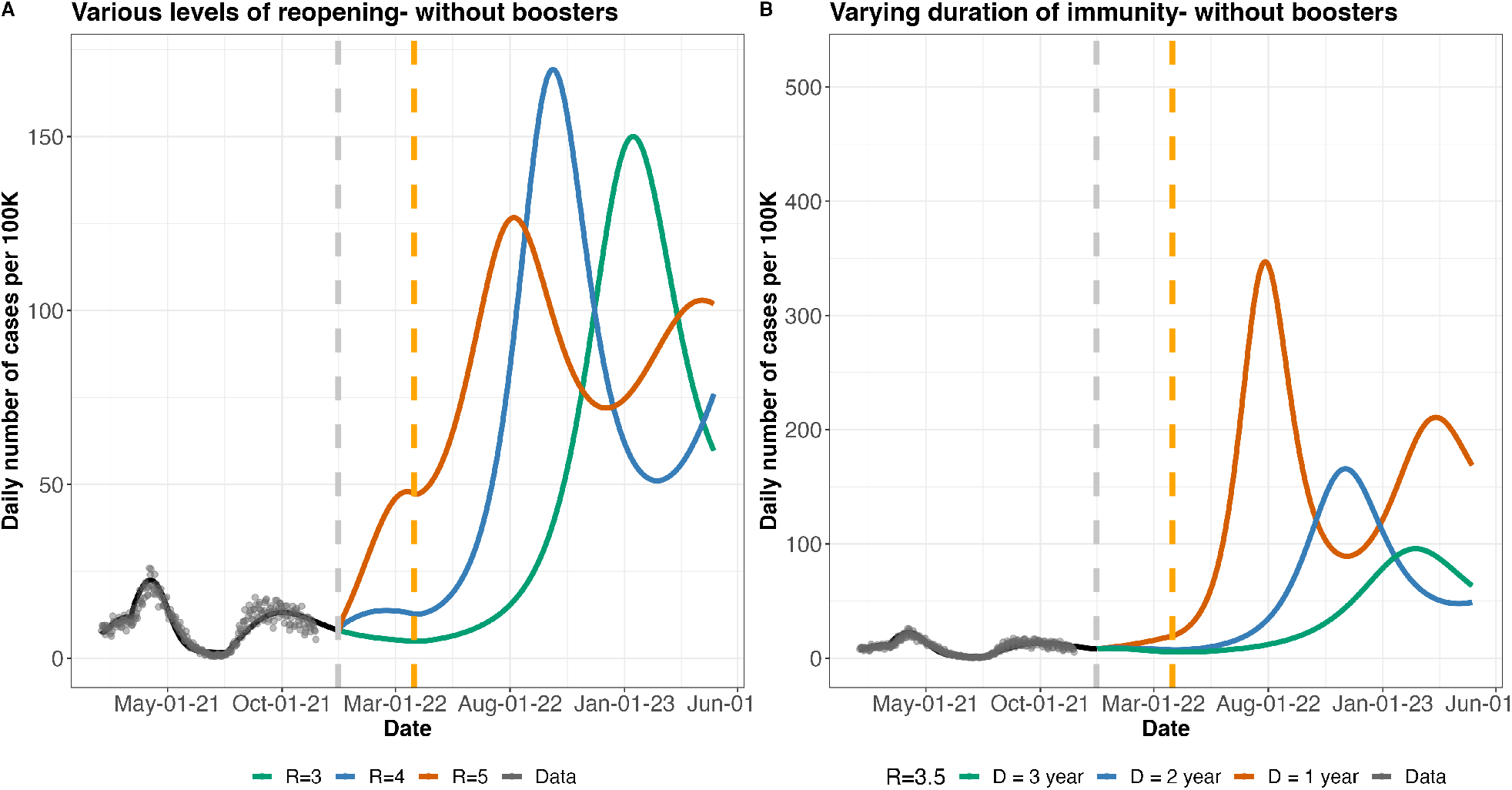
Near-future model projections and various possible paths to COVID-19 endemicity in BC without boosters. A. Projected daily cases for different levels of reopening: assuming the province reopen further at the end of November 2021. *R*_NPI_ indicates how much reopening is done. Importation rate *f* is fixed at 2 cases per 100K per day, vaccine efficacy *v*_*e*_ at 80%, and duration of immunity *D* is set to 2 years. B. Projected case numbers as a function of various lengths of duration of immunity (1 -3 years). While *R*_NPI_ is set to 3.5, f is 2 cases per 100K per day, *v*_*e*_ is 80%. The orange vertical dashed line indicate the point where vaccine boosters are suspended with 70% of the population completely immune to infection

**Figure S3:**
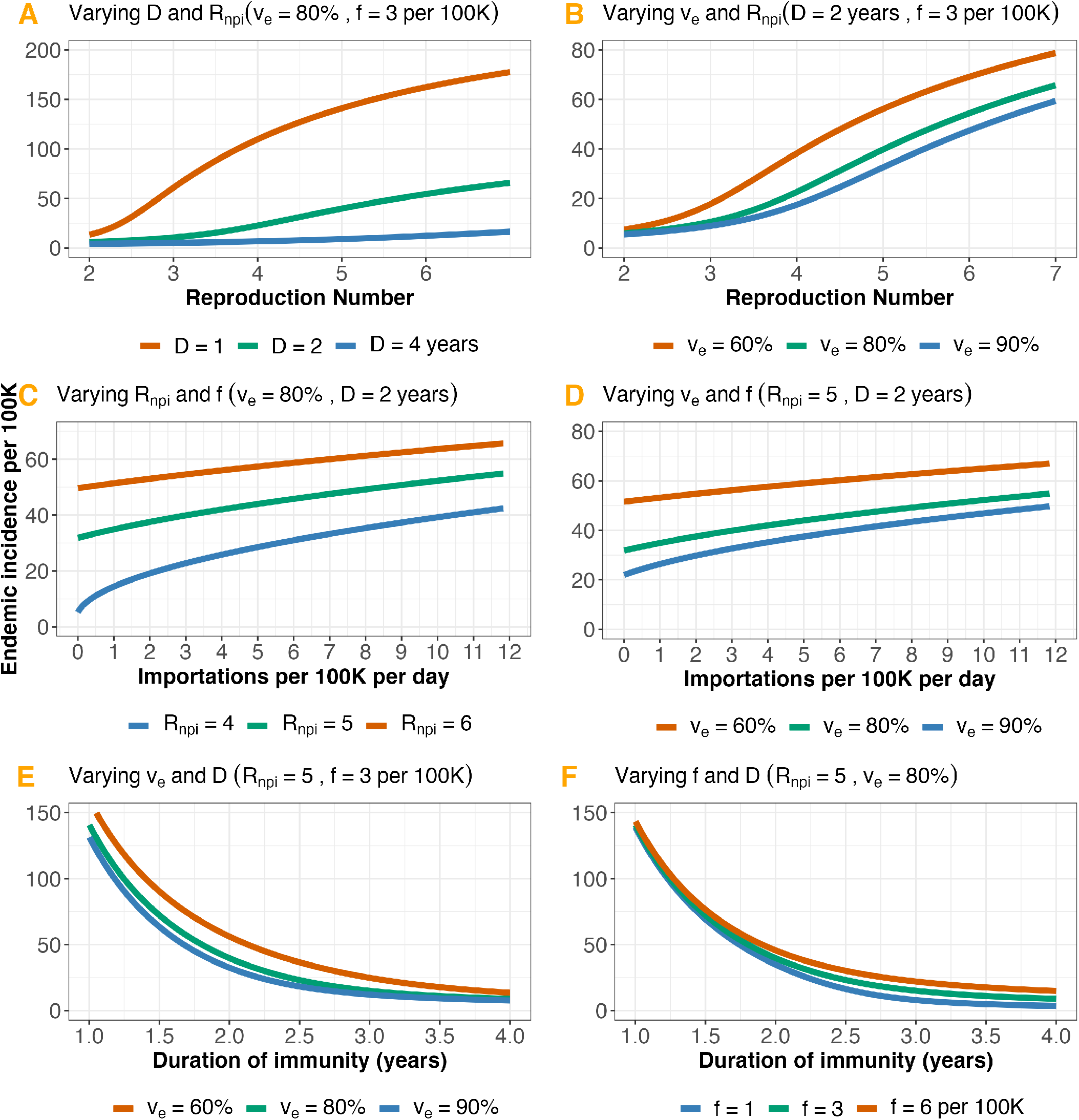
Endemic incidence as a function of: A. The reproduction number (*R*_NPI_) and duration of immunity (*D*). B. *R*_NPI_ and vaccine efficacy (*v*_*e*_). C. Importation rate (*f*) and *R*_NPI_. D. *f* and *v*_*e*_. E. *D* and *v*_*e*_. F. *D* and *f*. Baseline parameter values are: *f* = 3 cases per 100K per day, vaccination rate *ν* = 0.5% per day, *D* = 2 years, *v*_*e*_ = 80%, and *R*_NPI_ = 5.

**Figure S4:**
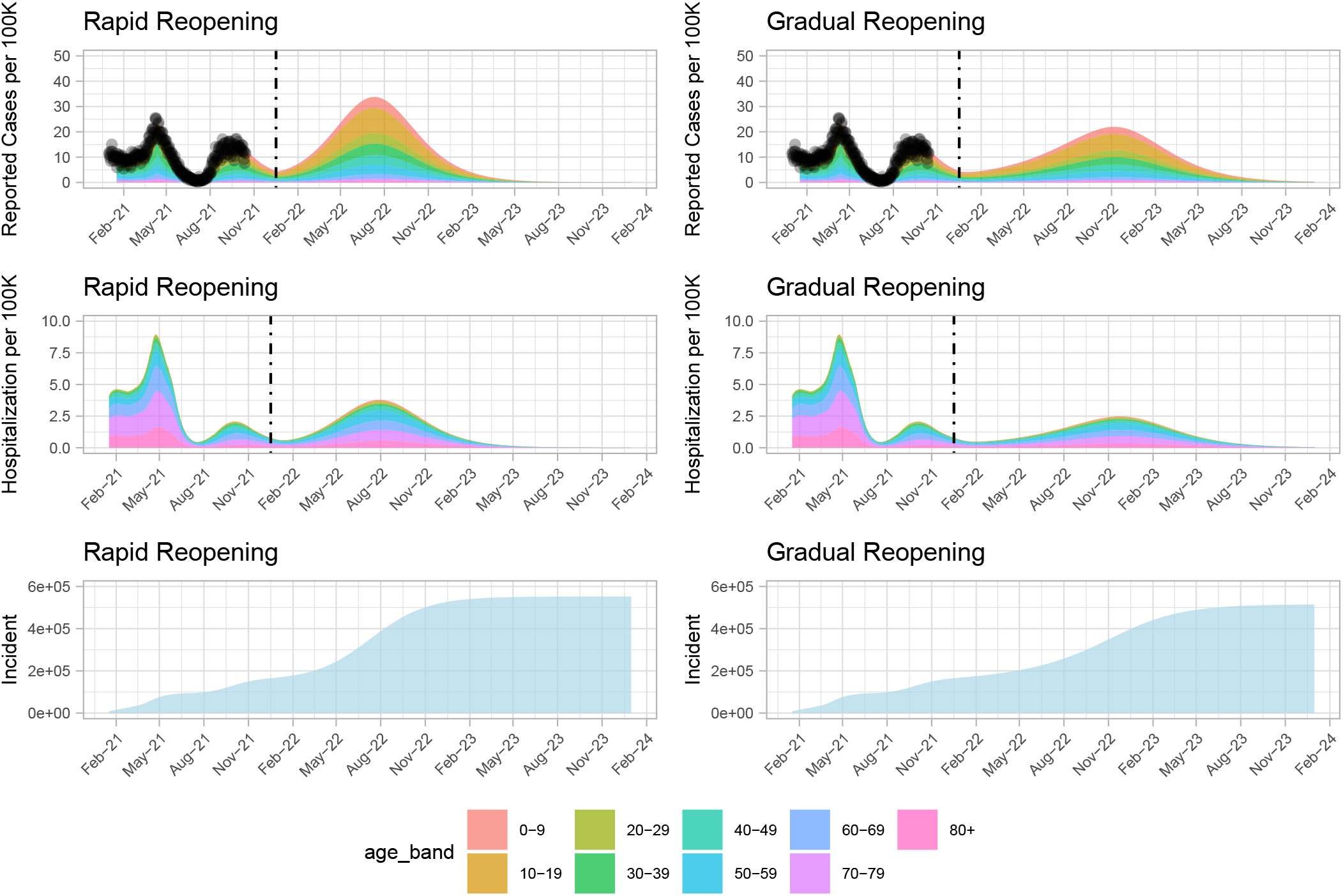
Comparison of the impact of gradual vs rapid reopening, from *R*_NPI_ = 2.2 to *R*_NPI_ = 2.4, on case and hospitalization rates. Gradual reopening is modelled as occurring over a 300 day window. Dashed lines represent reopening time. No further vaccines are deployed after reopening in both scenarios. The number of cases is 385K and 284K, for rapid and gradual scenarios, respectively, after reopening. The difference between scenarios equal 101K.The number of hospitalizations is 15K and 13K, for rapid and gradual scenarios, respectively, after reopening. The difference between scenarios equal 2K.

**Figure S5:**
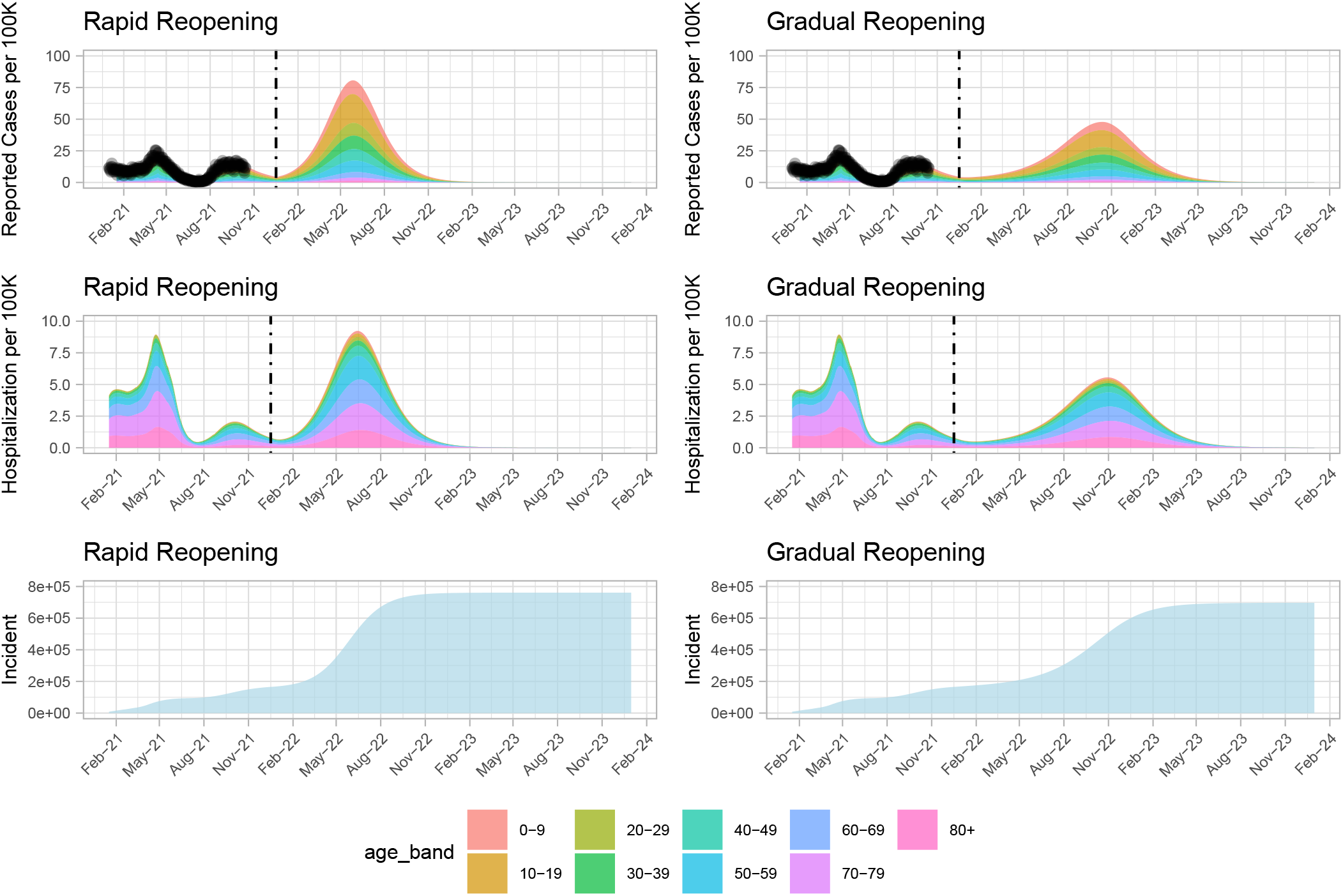
Comparison of the impact of gradual vs rapid reopening, from *R*_NPI_ = 2.2 to *R*_NPI_ = 2.6, on case and hospitalization rates. Gradual reopening is modelled as occurring over a 300 day window. Dashed lines represent reopening time. No further vaccines are deployed after reopening in both scenarios. The number of cases is 593K and 414K, for rapid and gradual scenarios, respectively, after reopening. The difference between scenarios equal 179K.The number of hospitalizations is 23K and 20K, for rapid and gradual scenarios, respectively, after reopening. The difference between scenarios equal 3K.

**Figure S6:**
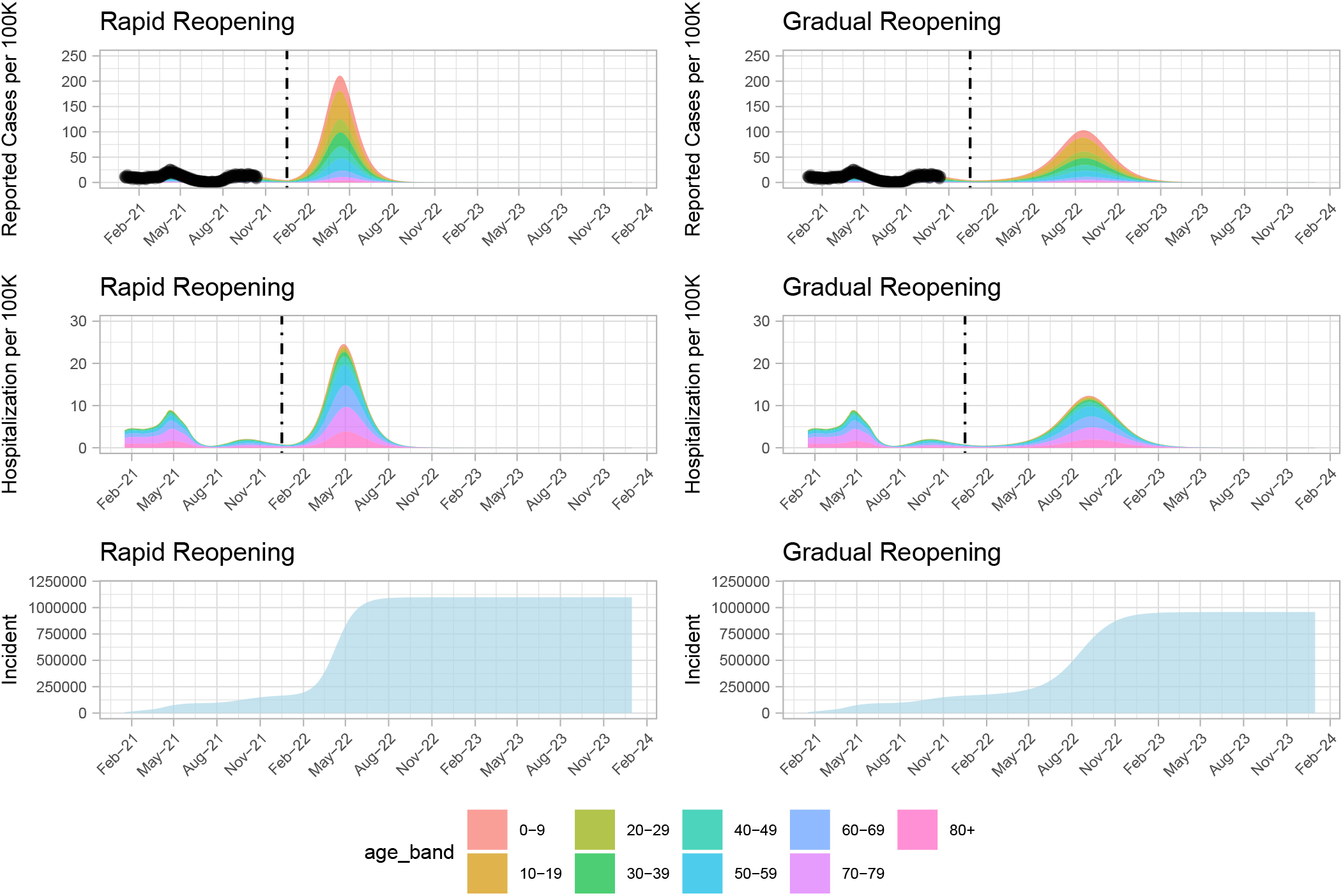
Comparison of the impact of gradual vs rapid reopening, from *R*_NPI_ = 2.2 to *R*_NPI_ = 3.0, on case and hospitalization rates. Gradual reopening is modelled as occurring over a 300 day window. Dashed lines represent reopening time. No further vaccines are deployed after reopening in both scenarios. The number of cases is 930K and 597K, for rapid and gradual scenarios, respectively, after reopening. The difference between scenarios equal 333K.The number of hospitalizations is 39K and 32K, for rapid and gradual scenarios, respectively, after reopening. The difference between scenarios equal 7K.

**Figure S7:**
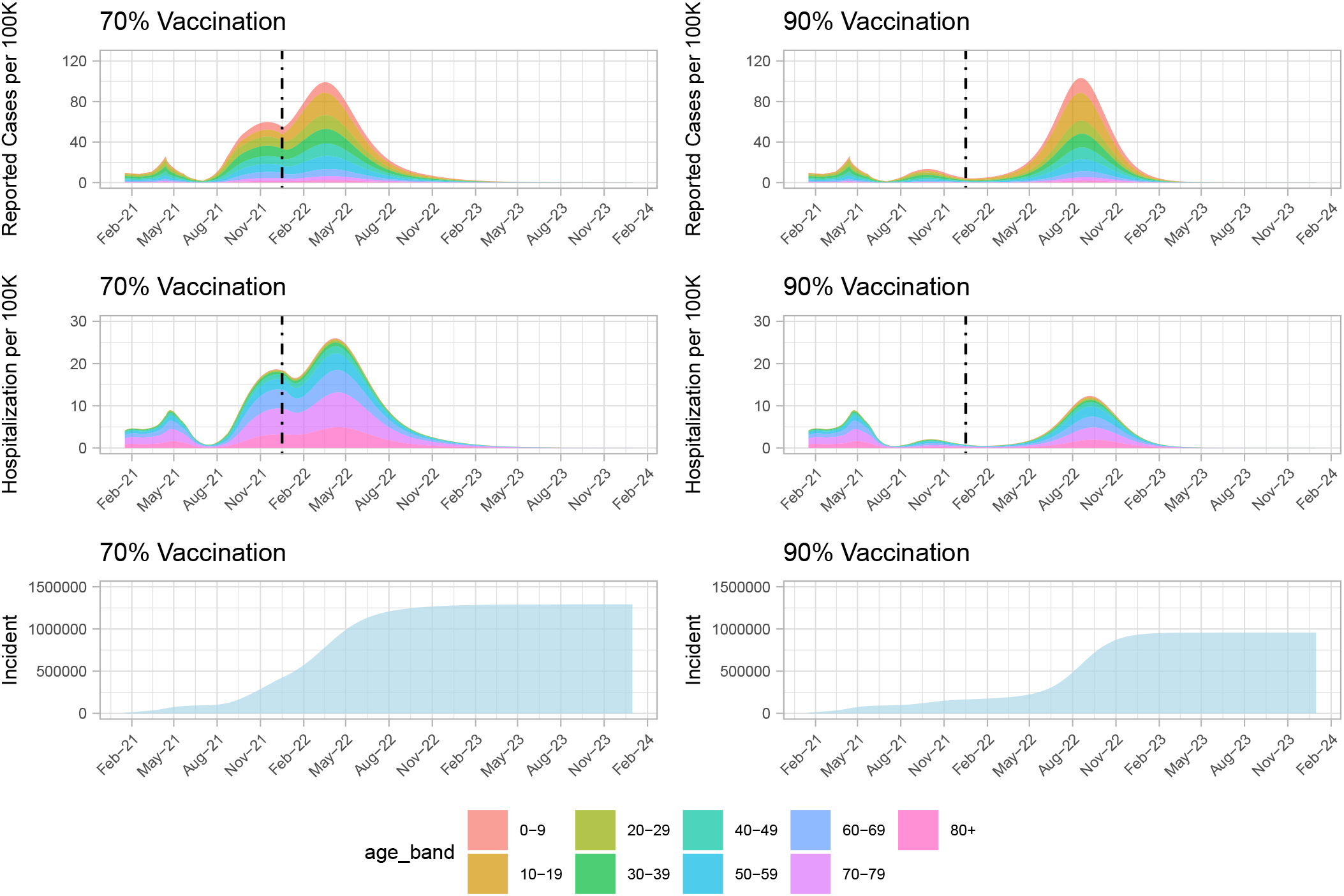
Reopen gradually at 70% or 90% vaccination coverage. Top panel showing reported cases per 100K of the population for the two vaccination coverage scenarios. Middle panel showing hospitalization per 100K of the population. Bottom panel showing the cumulative cases for the two vaccination coverage levels. Gradual reopening is modelled as occurring over a 300 day window. Dashed lines represent reopening time, from *R*_NPI_ = 2.2 to *R*_NPI_ = 3.0

**Figure S8:**
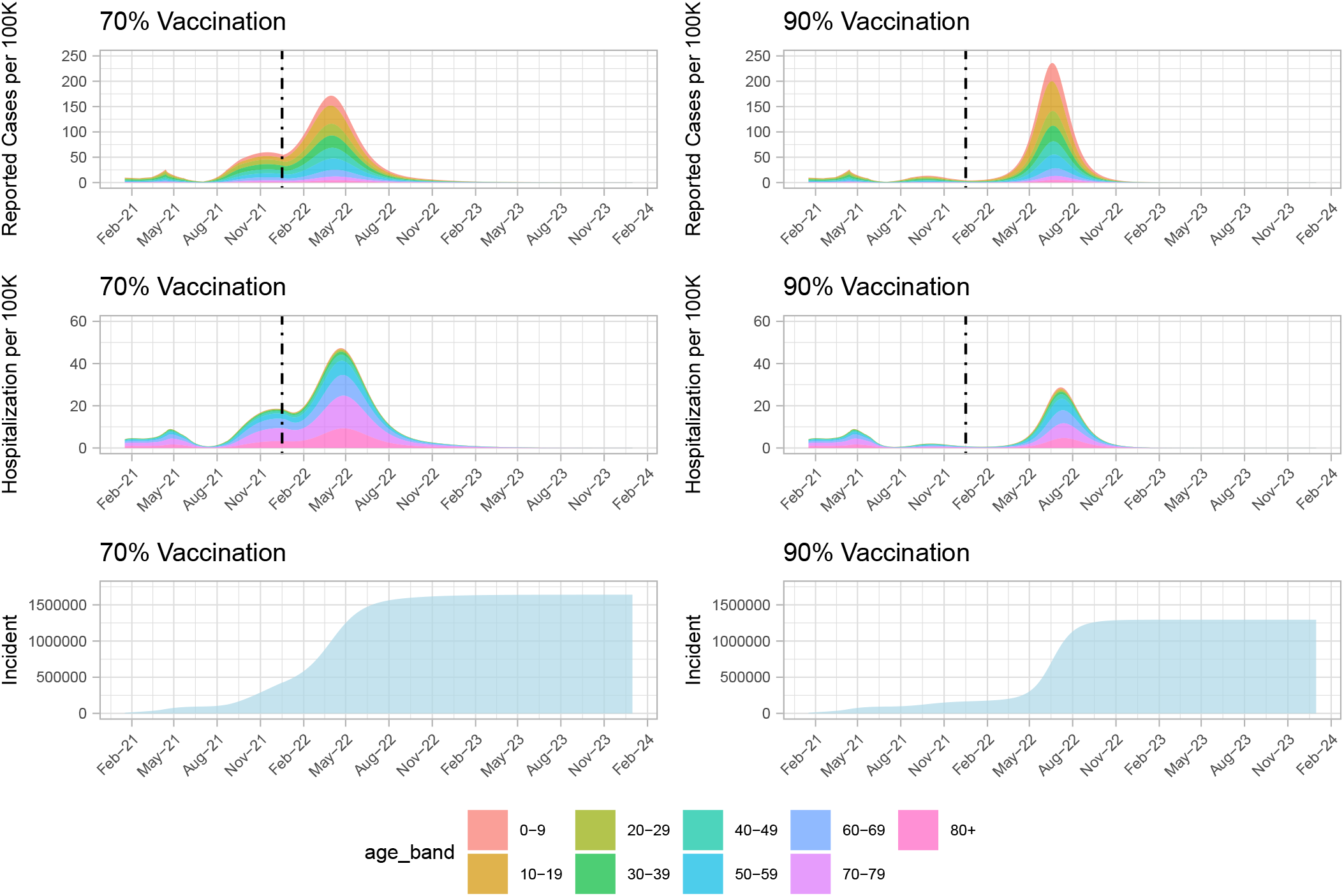
Reopen gradually at 70% or 90% vaccination coverage. Top frame showing reported cases per 100K of the population for the two vaccination coverage scenarios. Middle panel showing hospitalization per 100K of the population. Bottom panel showing the cumulative cases for the two vaccination coverage levels. Gradual reopening is modelled as occurring over a 300 day window. Dashed lines represent reopening time, from *R*_NPI_ = 2.2 to *R*_NPI_ = 4.0

**Figure S9:**
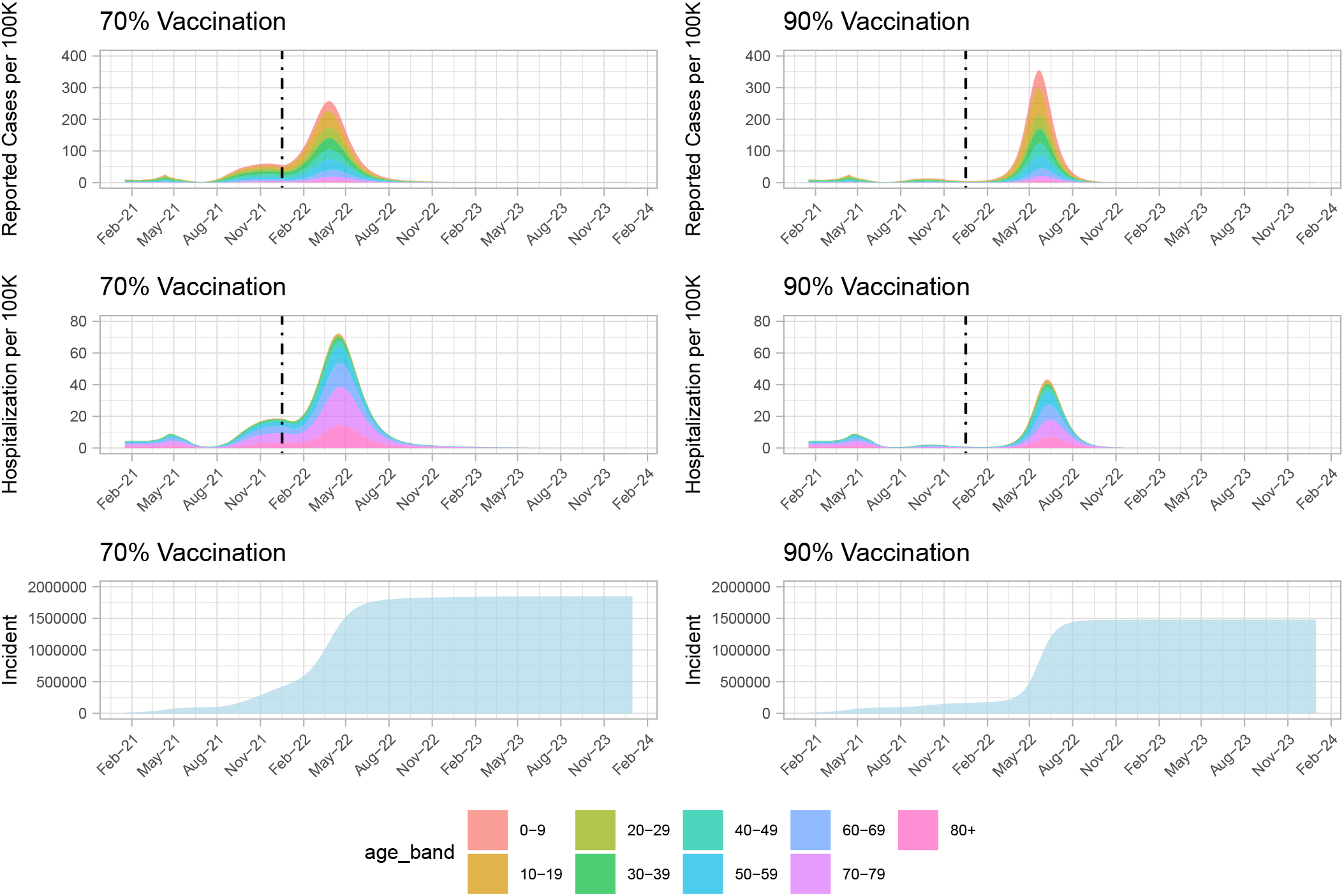
Reopen gradually at 70% or 90% vaccination coverage. Top frame showing reported cases per 100K of the population for the two vaccination coverage scenarios. Middle panel showing hospitalization per 100K of the population. Bottom panel showing the cumulative cases for the two vaccination coverage levels. Gradual reopening is modelled as occurring over a 300 day window. Dashed lines represent reopening time, from *R*_NPI_ = 2.2 to *R*_NPI_ = 5.0

**Figure S10:**
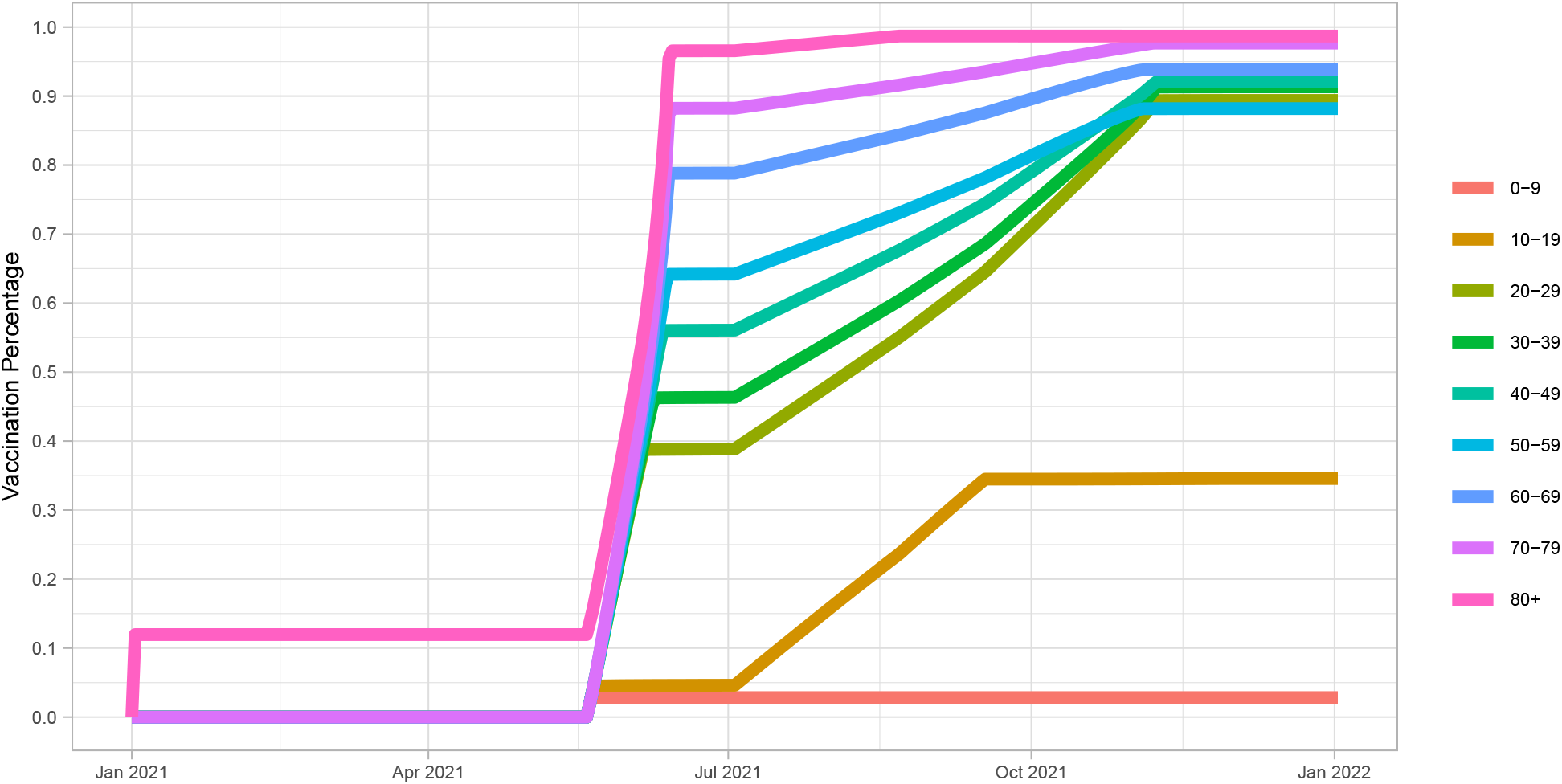
Vaccine rollout in the age and contact structured model. We assume 12% of the 80+ age group are fully vaccinated on January 01, 2021. Vaccination by age for those aged 20+ starts on May 22, 2021 and for those aged 10-19 starts on July 3,2021. This figure includes essential workers.

**Table S1:** Specifics of the age-based vaccine rollout in the age and contact structured model.

| Age group | Vaccination start date | May-June uptake | Overall uptake |
| --- | --- | --- | --- |
| 0-9 | No vaccination | 0% | 0% |
| 10-19 | 2021-07-03 | 0% | 35% of those aged 12-19 |
| 20-29 | 2021-05-22 | 40% | 89% |
| 30-39 | 2021-05-22 | 45% | 90% |
| 40-49 | 2021-05-22 | 55% | 92% |
| 50-59 | 2021-05-22 | 65% | 88% |
| 60-69 | 2021-05-22 | 80% | 95% |
| 70-79 | 2021-05-22 | 88% | 97% |
| 80+ | 2021-01-01 | 95% | 98% |

## Important Dates Specification

**Table S2:** Important dates were identified from British Columbia COVID-19 reports that illustrate rapid changes in daily cases. These important dates are determined in the table.

| Important Date | Description |
| --- | --- |
| 2021-01-01 | Simulation starts |
| 2021-02-15 | The number of cases gradually decreased to 400 |
| 2021-03-21 | The number of cases gradually increased to 575 |
| 2021-04-15 | The number of cases rapidly increased to 1250 |
| 2021-07-22 | The number of cases rapidly decreased to 45 |
| 2021-09-01 | The number of cases gradually increased to 500 |

## Notes

### Competing Interest Statement

The authors have declared no competing interest.

### Author Declarations

http://www.bccdc.ca/health-info/diseases-conditions/covid-19/data

